# CHARACTERIZATION OF THE PROPORTION OF CLUSTERED TUBERCULOSIS CASES IN GUATEMALA, CA: INSIGHTS FROM A MOLECULAR EPIDEMIOLOGY STUDY, 2010-2014

**DOI:** 10.1101/2020.03.23.20033878

**Authors:** María Eugenia Castellanos, Dalia Lau-Bonilla, Anneliese Moller, Eduardo Arathoon, Frederick D. Quinn, Mark H. Ebell, Kevin K. Dobbin, Blanca Samayoa, Christopher C. Whalen

## Abstract

**Background:** There is little information about the proportion of clustering of tuberculosis cases from low-income settings, which can represent ongoing transmission events. We investigated for the first time the proportion of clustered tuberculosis cases based on genotypic matching in Guatemala City, Guatemala between 2010 and 2014 and potential risk factors associated with these clustered cases in HIV-infected subjects. Moreover, the genetic diversity of *M. tuberculosis* isolates in this country is presented.

**Design and methods:** This study was a retrospective observational study conducted on *Mycobacterium tuberculosis* isolates from HIV-infected and non-HIV infected tuberculosis cases that submitted samples to a referral tuberculosis laboratory in Guatemala City, Guatemala from 2010-2014. Genotyping results were compared with the international spoligotyping database, SITVIT2 and classified accordingly. We generated a spoligoforest using the MERCAT program. We categorized spoligotype patterns as clustered or non-clustered depending of their genotype and estimated the proportion of clustering and the recent transmission index (RTI_n-1_). We analyzed the crude association between demographic, clinical and behavioral variables and clustering in the HIV-population.

**Results:** From 2010 to 2014, a total of 479 patients were confirmed as tuberculosis cases by culture at the study site. Spoligotype patterns were available from 391 patients (82%), nine of them with two isolates included in the study. We detected 71 spoligotype patterns and overall, the most frequent spoligotyping families were LAM (39%), followed by T (22%), Haarlem (14%), X (13%), Unknown (6%) and Beijing (3%), representing 97% of the isolates. Out of the 400 isolates, 365 (91%) were grouped in 36 clusters (range: 2-92). The recent transmission index (RTI_n-1_) was 82%. Pulmonary tuberculosis was strongly associated with clustering in the 113 HIV-infected group with available data (OR=4.3, 95% CI 1.0-17.7).

**Conclusion:** There might be high levels of ongoing transmission of *M. tuberculosis* in Guatemala City, Guatemala as indicated by clustering in a convenience sample. Among HIV-infected patients, clustering was more likely in pulmonary disease.

## INTRODUCTION

*Mycobacterium tuberculosis* strains are considered clustered when the genotypes between two or more are the same. Clustered strains represent a chain of transmission and may represent ongoing or recent transmission if the initial isolates are collected during a short period of time, such as one to two years, in a defined geographic area. Thus, when the sample collection interval is narrow enough, and in a well-defined geographical area [1], we can infer that clustered cases represent recent transmission; the assumption being that clusters are “epidemiologically linked chains of recently transmitted disease” [2]. Strains with unique genotypes are thought to represent reactivation of an old tuberculosis infection and are considered non-clustered.

The distribution of clustered strains is affected by several host and population-level characteristics [3, 4]. Individual host characteristics that influence levels of tuberculosis clustering include place of birth, pulmonary tuberculosis disease (rather than extrapulmonary), and alcohol abuse [4]. A few population-level characteristics that affect clustering are age structure of the population, prevalence of latent tuberculosis infection and HIV prevalence [3]. Several studies have been performed evaluating levels of tuberculosis clustering and potential risk factors for increased cluster risk. Most of these studies, however, were conducted in low-incidence tuberculosis settings and little is known about relevant risk factors for recent transmission in HIV-infected patients in areas with a medium tuberculosis burden, such as Guatemala.

The main aim of this study was to characterize the proportion of clustered tuberculosis cases based on genotypic matching in Guatemala City, Guatemala between 2010 and 2014 and to identify risk factors associated with these clustered cases in HIV-infected subjects. As a secondary aim, we present for the first time the genetic diversity of *M. tuberculosis* isolates from patients in Guatemala. The international genotyping database SITVIT2, which contains data from 111,635 isolates from 169 countries, was employed to classify and relate the Guatemalan strains to the global framework of tuberculosis [5]. Knowledge of existing circulating genotypes and the identification of risk factors associated with recent transmission will allow an evidenced-based approach for health policymakers to direct and concentrate targeted tuberculosis control measures to high-risk populations

## STUDY POPULATION AND METHODS

### DESIGN OVERVIEW

This is a retrospective observational study. Genotypes of *M. tuberculosis* isolates from HIV-infected and non-HIV infected tuberculosis cases in Guatemala City from 2010-2014 were categorized as clustered or non-clustered depending of their genotype. Clustered strains were considered as evidence of recent transmission. Independent potential factors associated with having a clustered isolate were investigated in the HIV-infected subjects.

### STUDY POPULATION, SETTING AND DATA SOURCES

#### Study Setting

The study was conducted in Guatemala City, Guatemala at Integral Health Association (ASI). In Guatemala, in 2014, 4,200 new tuberculosis cases were notified for an incidence rate of 25 cases per 100,000/year [6]. However, the World Health Organization has estimated that due to the low case detection rate, the incidence could be almost two times higher, with an estimated incidence of 60 cases/100,000/year. Out of these 4,200 individuals with incident TB, 270 were HIV infected subjects.

ASI is an institution which has served over 25 years in Guatemala City, and has supported particularly the management of HIV-infected subjects. ASI operates a laboratory that serves as a referral center for the diagnosis of tuberculosis and other infectious diseases, receiving samples from Guatemala City and other regions of the country.

#### Study population

Patients with tuberculosis who submitted samples for the diagnosis of tuberculosis at the laboratory of ASI in Guatemala City during 2010-2014 were included in this study.

#### Inclusion and exclusion criteria

We only included individuals in whom a *M. tuberculosis* isolate was detected during 2010-2014. *Mycobacterium tuberculosis* isolates should have been confirmed as such by laboratory methods (culture plus specie identification by conventional or molecular methods). Isolates without a clinical record nor a spoligotyping results were excluded. We also excluded isolates with multiple spoligotyping results per isolate but without at least two identical results per sample.

#### Definition of HIV status in the study population

Patients managed and treated in an urban HIV clinic who continually submits samples to the laboratory of ASI were considered HIV-infected individuals. Other patients were considered as HIV negative/unknown status.

#### Genotyping technique

Isolates were analyzed by spoligotyping, as described in detail elsewhere [7]. Briefly, direct repeats within the *M. tuberculosis* genome are interspersed with polymorphic DNA sequences called “spacers”. In this technique, both the direct repeats and the spacers are amplified by polymerase chain reaction (PCR). Oligonucleotides that correspond to 43 of these spacers are immobilized into a membrane, and the amplified fragments are hybridized to these spacers. The hybridization products are detected by chemiluminescence [7], and depending on the presence and distribution of the spacers, a genotype pattern is obtained for each isolate.

#### Data source

Data regarding the samples (time of tuberculosis diagnosis, type of sample, spoligotyping results and drug susceptibility of the isolates) and the HIV-infected individuals (demographic, clinical and behavioral data) were extracted from the records submitted to ASI.

### STUDY OUTCOME FOR CLUSTERING

The study outcome was the clustering of a strain. A clustered strain was defined as one that at least share the same genotype with another isolate, regardless of the HIV status of the corresponding patients. Isolates with unique genotype patterns were considered non-clustered/unique. Strains that belong to any cluster were coded as one (1) and strains that do not share a genotype pattern with any other isolate were coded as zero (0).

### STUDY EXPOSURES FOR CLUSTERING

In the case of HIV-infected patients, an extensive set of demographics, clinical and behavioral variables were evaluated as independent potential factors for clustering. The complete list and categorization of these variables are presented in Table S1.

### ANALYTICAL STRATEGY

#### Data preparation and cleaning

From the main analysis, we excluded all isolates without spoligotyping results. When an isolate had multiple spoligotyping results, we kept the spoligotype that had at least two identical results. For each patient, the first available sample was selected. We excluded patients with different spoligotypes if the strains were isolated in a time period of less than six months and the samples came from the same source (e.g. pulmonary versus extra-pulmonary). If the time period was more than six months or if the samples came from different source (pulmonary versus extra-pulmonary), we considered them as distinct patients regardless of the spoligotyping result. After the cleaning, each patient had a unique spoligotype, and the source of the sample(s) was categorized as pulmonary or extra-pulmonary. Patients with pulmonary and extra-pulmonary samples collected at the same time, were considered as extra-pulmonary cases.

We transformed the octal format of the spoligotypes to binary format. This format should contain 43 digits, representing the presence (1) or absence (0) of 43 spacers, but for technical issues at the study site only 42 digits were captured. We corrected the data using SpolSimilaritySearch, an online tool with a collection of over 100,000 isolates from 169 countries of origin of the tuberculosis patients (http://www.pasteur-guadeloupe.fr:8081/SpolSimilaritySearch/index.jsp). We assigned the most frequent last spacer that was found in the SpolSimilaritySearch, database (the selected spacer should have a frequency of over 95% in the database) to obtain the corrected spoligopattern (Appendix file). Patterns where the frequency of the last spacer in the SpolSimilaritySearch, database was <95% for either their absence of their presence, and unique patterns that have never been reported were excluded of the analysis as we could not assign the last spacer.

#### Study population

We estimated the proportion of tuberculosis cases with spologytping results out of the total number of patients with a positive *M. tuberculosis* culture. For all patients, we estimated the type of sample source (pulmonary and extrapulmonary) and the drug susceptibility patterns of their isolates (resistant or non resistant) using proportions. We stratified these results by HIV status of the patients.

#### Assignment of shared international type and spoligotyping families

We assigned each spoligotype a shared international type (SIT) and family using the international database SITVIT2, and update of SITVITWEB (http://www.pasteur-guadeloupe.fr:8081/SITVIT2/) [5]. If there was no SIT listed, but the pattern was reported in the database as an orphan isolate, we named the pattern as ‘Pseudo-XX’ whereby the XX represents a two-digit number that we assigned. We estimated the frequency of these SITs and families using proportions, in the overall population and per year of isolation.

#### Characteristics of the patients with two samples included in the study

We described the major characteristics of the patients that provided two samples for the study: HIV status, sex, age at time of tuberculosis diagnosis, type of sample, time between tuberculosis diagnosis, acid-fast staining result, drug susceptibility pattern, SIT, spoligotyping family, CD4 count, previous history of tuberculosis, behavioral risk factors for clustering, and record of death.

#### Visualization of relationship among spoligotypes

We constructed a spoligoforest, a visualization of relationships among spoligotypes [8], based on mutation events, using the new computational open-source package MERCAT-Molecular Epidemiology Researcher’s Collection of Analytical tool- [9], an expansion of the online tool “SpoolTools” [10]. In this type of graph, each node represents a spoligotype. The size of each node represents the number of strains that belongs to a given spoligotype. Each node is labelled with consecutive numbers, starting with the node with the highest number of samples. Directed edges indicate single-event deletion that relates to two clusters, the arrowheads pointing to the descendants. The spoligoforest was colored by spoligotyping family using the open source graph visualization software Inkscape (https://inkscape.org/). We used the same colors that were used from the authors of the SITVIT2 in the most recent report [5].

#### Descriptive statistics of clustering of mycobacterial isolates

Descriptive statistics were used to present the number and proportion of clustered strains and clusters, distribution of cluster size, mean size of cluster, maximum size of cluster, proportion of cluster with 2 cases, clusters 3-19 cases and clusters ≥20 cases. We selected these statistics based on previous literature [4, 11-14].

#### Estimation of the proportion of TB cases due recent transmission

Several methods to estimate the proportion of tuberculosis cases due to recent transmission have been described [12]. We used the “n-1” method, applying the original formula, developed by Small and others: Recent Transmission Index, RTI_n−1_ = (n_c_ − c)/n [15], in which n = total number of cases in the sample, c = is the number of clusters (genotypes represented by at least two cases) and n_c_ =is the total number of cases in a cluster of two or more. As suggested by Glynn and colleagues [16] because of the length of this study, we re-estimated the proportion of tuberculosis cases due to recent transmission using different time windows: 2 years (2010-2011), 3 years (2010-2012) and 4 years (2010-2013).

#### Independent factors associated with clustering in the HIV-infected population

Potential risk factors for clustering (previously described in “Study exposures for clustering” section) were reported for the HIV-population and were compared by Chi-square test (categorical variables), Fisher Test (counts less than 5) or Wilconxon test (continuous variables) in patients with clustered and non-clustered strains. In the categorical variables with more than two classes and when the overall *p* values were less than 0.20, a Bonferroni correction was conducted: A corrected *p* value was obtained for the pairwise comparison between a given class and the reference class.

In the variables and classes in which *p* values < 0.20 were obtained, we estimated the crude association between each of these predictors and clustering using regression models. In these regression models, the outcome, or dependent variable, is clustering as defined in section “Study Outcome for Clustering”. Since this is a dichotomous outcome variable, at logistic model was initially considered appropriate. However, our sample size was small and there was a very low proportion of one of the events (non-clustered strains). Thus, we estimated the association between exposure and the outcome using the Firth logistic regression method [17]. Unadjusted odds ratio and prevalence ratio (with 95% CI) were obtained with this method by exponentiating the regression coefficients. Due to the low proportion of non-clustered strains we did not conduct multivariate regression models.

All statistical analyses were conducted on SAS software (release 9.4, SAS Institute Inc., Cary, NC, USA) and R v3.6.0 (R Foundation for Statistical Computing, Vienna, Austria, 2019).

### SENSITIVITY ANALYSIS

We re-analyzed the proportion of clustering and RTI_n−1,_ after exclusion of clusters with a large size, ≥20 isolates. This approach has shown to improve at least partly the specificity of spoligotyping [18].

### ETHICAL APPROVAL

Institutional review board clearance was obtained from Zugueme, a Guatemalan independent Ethics Committee, approved by the Ministry of Health of Guatemala and by the University of Georgia.

## RESULTS

### Study population

From 2010 to 2014, a total of 479 patients were confirmed as tuberculosis cases by culture at the study site. Three hundred ninety-one (82%) had isolates with valid spoligotyping results. Of these, there were 7 patients who provided two samples collected at different time points (> 6 months apart). In addition, there were 2 patients who presented mixed infections, with samples collected concurrently from different sites (pulmonary and extra-pulmonary) with different spoligotype pattern (later described in detail).

Based on our methodology, we considered these 9 cases as 18 patients, thus the final sample size of the study was 400 patients (Figure 1). Of these patients, 132 were confirmed as HIV-infected (33%). Most patients provided pulmonary samples (68%). Overall, 32% of the patients had strains resistant to at least one of the anti-tuberculosis drugs and 1% of them were multi-drug resistant (MDR) (Table 1).

**Table 1.** Type of sample and drug resistance pattern of *M. tuberculosis* isolates from included patients, Guatemala City, Guatemala from 2010-2014. Results are shown for the overall population and stratified by HIV status.

| Characteristic | All<br>N=400 |  | HIV infected<br>n=132 |  | HIV status<br>negative/unknown<br>n=268 |  |
| --- | --- | --- | --- | --- | --- | --- |
|  | N | % | N | % | N | % |
| Type of sample |  |  |  |  |  |  |
| Pulmonary | 270 | 68 | 92 | 70 | 178 | 66 |
| Extrapulmonary | 130 | 33 | 40 | 30 | 90 | 34 |
| Isoniazid (n=300) |  |  |  |  |  |  |
| Sensitive | 259 | 86 | 112 | 93 | 147 | 82 |
| Resistant | 41 | 14 | 8 | 7 | 33 | 18 |
| Rifampicin (n=301) |  |  |  |  |  |  |
| Sensitive | 289 | 96 | 114 | 94 | 175 | 97 |
| Resistant | 12 | 4 | 7 | 6 | 5 | 3 |
| Pyrazinamide (n=102) |  |  |  |  |  |  |
| Sensitive | 96 | 94 | 81 | 94 | 15 | 94 |
| Resistant | 6 | 6 | 5 | 6 | 1 | 6 |
| Streptomycin (n=301) |  |  |  |  |  |  |
| Sensitive | 234 | 78 | 99 | 82 | 135 | 75 |
| Resistant | 67 | 22 | 22 | 18 | 45 | 25 |
| Ethambutol (n=301) |  |  |  |  |  |  |
| Sensitive | 288 | 96 | 115 | 95 | 173 | 96 |
| Resistant | 13 | 4 | 6 | 5 | 7 | 4 |
| Any drug resistance (n=301) |  |  |  |  |  |  |
| No | 205 | 68 | 85 | 70 | 120 | 67 |
| Yes | 96 | 32 | 36 | 30 | 60 | 33 |
| MDR† resistance (n=300) |  |  |  |  |  |  |
| No | 296 | 91 | 118 | 98 | 178 | 99 |
| Yes | 4 | 1 | 2 | 2 | 2 | 1 |

†MDR=Multidrug-resistant, defined as resistant to at least isoniazid and rifampicin

**Figure 1.**
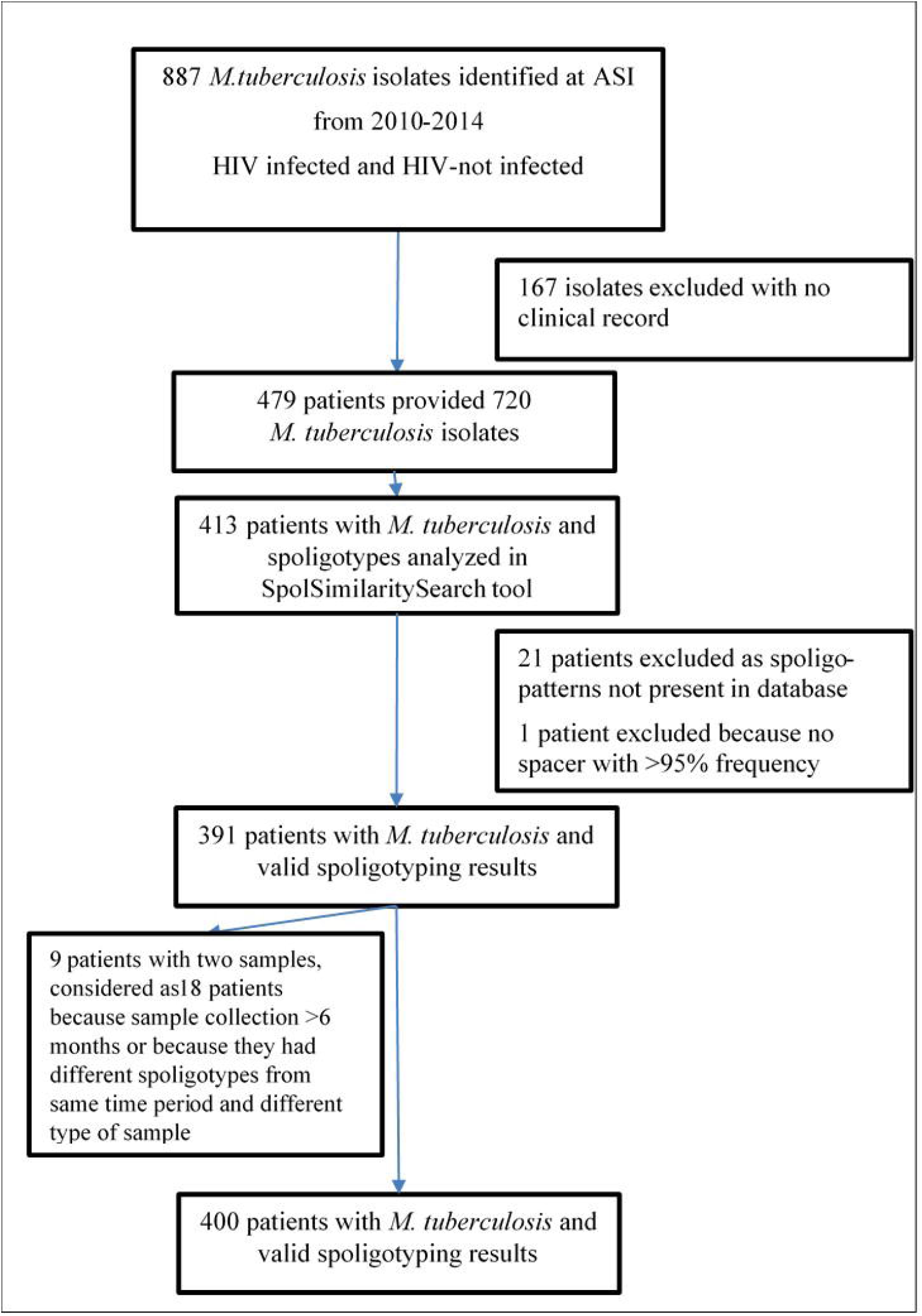
Flow diagram of the study. From 2010-2014, 887 *M. tuberculosis* isolates were identified at the tuberculosis laboratory at ASI, Guatemala City, Guatemala. Out of these, 720 *M. tuberculosis* isolates belonged to 479 tuberculosis cases. After data cleaning, we included in this study 400 tuberculosis cases with a valid spoligotyping result and estimated the proportion of clustering cases.

### Spoligotype patterns and families

We detected 71 spoligotype patterns, with 13 genotypes being present in ≥ 6 samples and comprising 75% of the total samples (Table 2). The five most frequent genotypes were SIT 33 (23%) from the Latin American-Mediterranean (LAM) family, SIT 53 (14%) from the ill-defined family T, SIT 42 (7%) from the LAM family, SIT 50 (6%) from the Haarlem family, and SIT 119 (5%) from the X family. The complete list of the 71 spoligotypes identified among the 400 cases is presented in Supplemental material (Table S2). Overall, the most frequent spoligotyping families were LAM (n=156, 39%), followed by T (n=89, 22%), Haarlem (n=57, 14%), X (n=51, 13%), Unknown (23, 6%) and Beijing (n=13, 3%), representing 97% of the isolates. The Ural, East African-Indian, Bovis, MANU and S families were uncommon (5 or less isolates). The frequency of the most common spoligotyping families did not vary substantially by year of isolation (Figure 2).

**Table 2.**
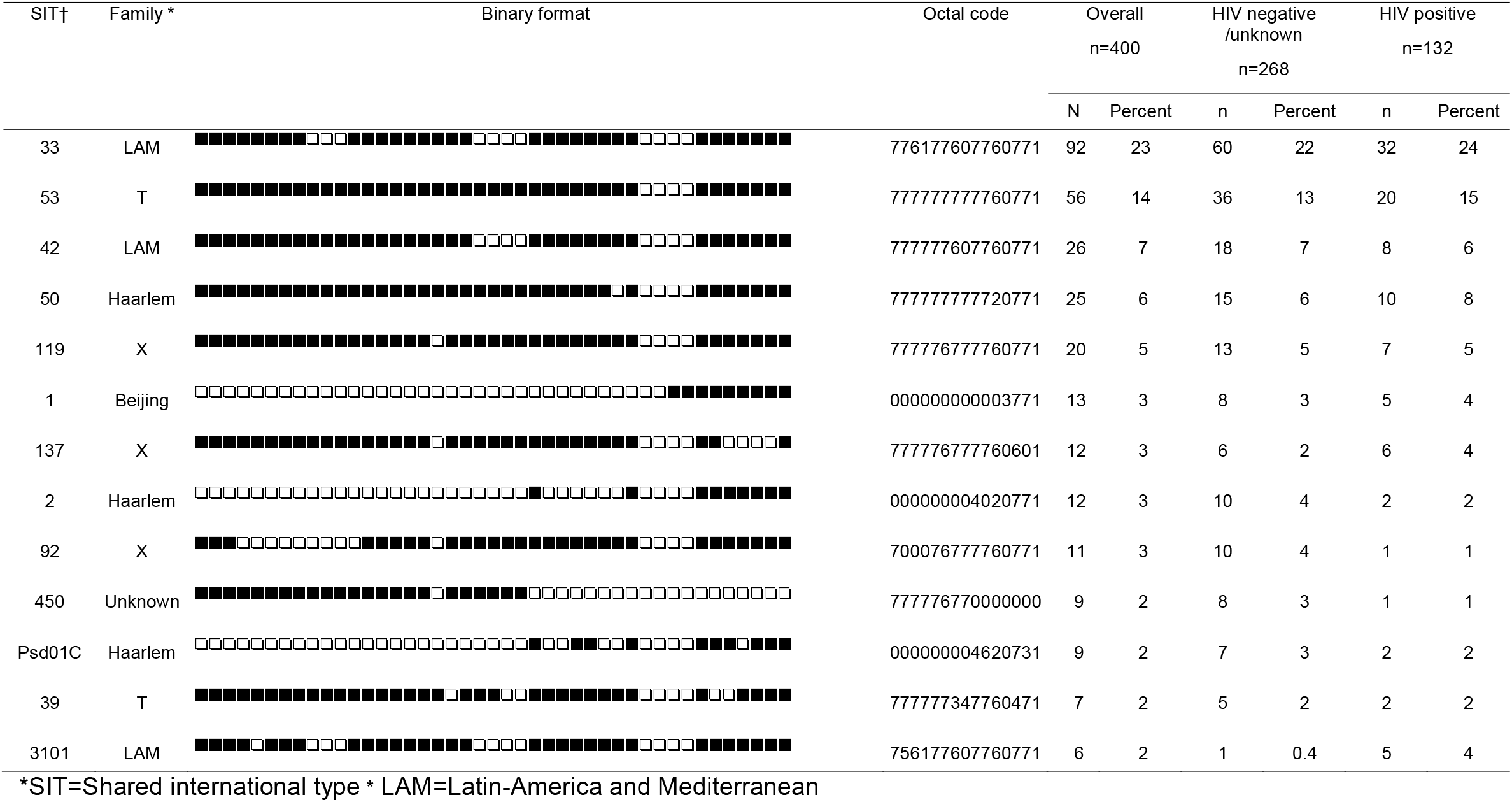
Most frequent spoligotypes of *M. tuberculosis* strains from tuberculosis cases in Guatemala City, Guatemala. Distribution of the 13 most frequent spoligotypes identified in the study. For each spoligotype, the shared international type (SIT), spoligotype family, binary format, octal code and frequency are presented. Results are shown for the overall population and stratified by HIV status. These 13 genotypes comprised 75% of the total samples.

**Figure 2.**
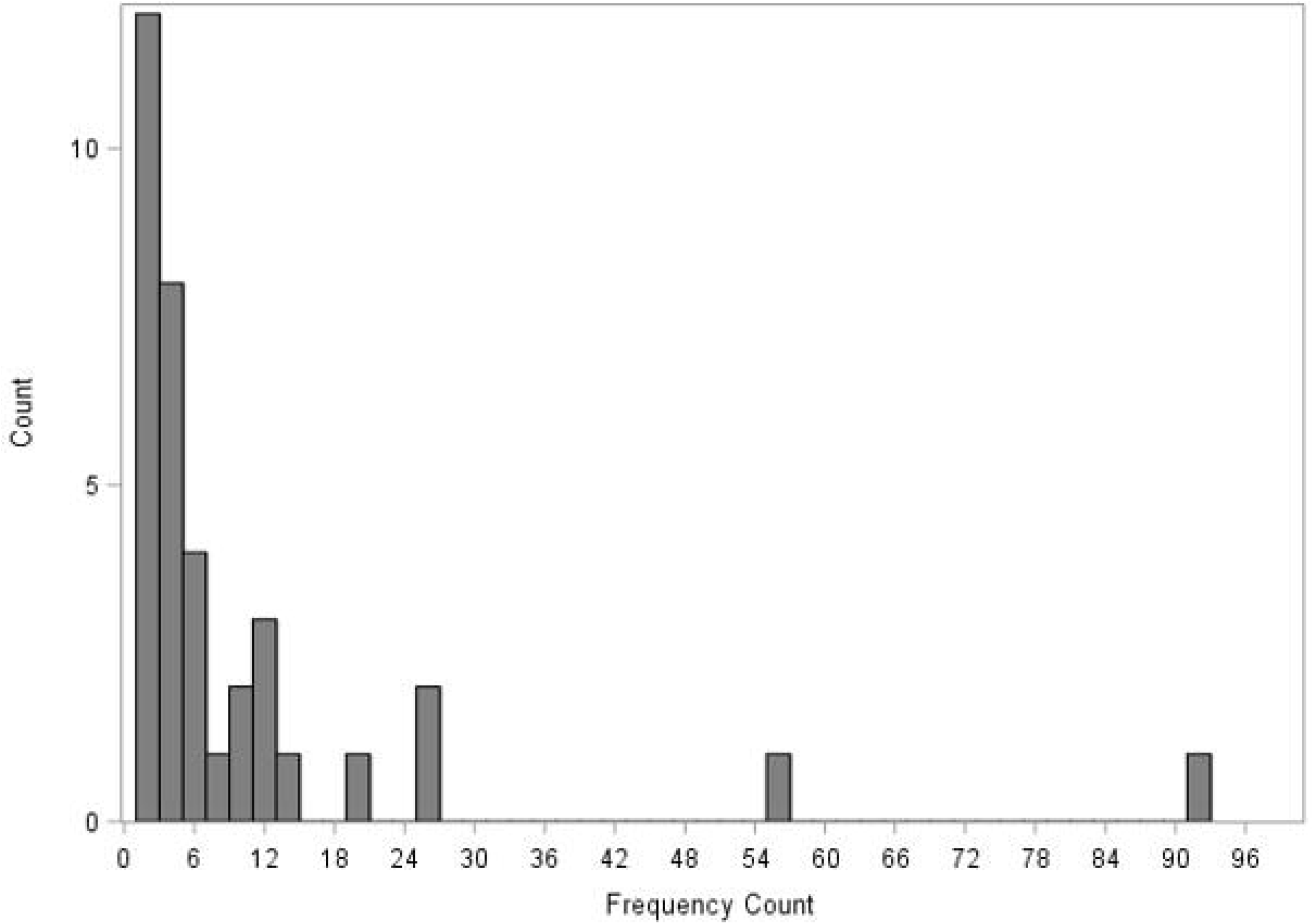
Spoligotypes lineages from *M. tuberculosis* isolates of included patients, Guatemala City, Guatemala, 2010-2014. Each bar is divided into colored segments indicating the relative percentage of each spoligotyping family detected each year.

### Characteristics of the nine patients with two samples

In the case of the two patients with mixed infections, their two samples presented different spoligotyping families. For study ID 97, the sputum sample had a spoligotype belonging to the Haarlem family, whereas the bone marrow sample had a spoligotype from the X family. For study ID 203, the sputum sample had a LAM family, whereas the stool sample had a spoligotype of Unknown family (Table S3). The latter patient had a co-infection with HIV, with a CD4 count of 28 cells/mm^3^. In the seven patients with two tuberculosis episodes with a difference of > 6 months, six of them presented the same spoligotype in the different time points, whereas only in one case (study ID 334) the samples had different spoligotype patterns (SIT 53, T family and SIT 42, LAM family); she was a woman that had been in prison. Also, it is worth to note that eight of these nine patients were HIV-infected individuals. A comprehensive description of the major characteristics of these patients and their samples are presented in Table S3.

### Visualization of relationship among spoligotypes

Based on the results of the spoligoforest (Figure S1) SIT 53 (node 2) was the root node and the most likely oldest spoligotype. SIT 42 (node 3) was the precursor of the biggest cluster in the study (SIT 33, node 1). These three SITs, 53, 33 and 42 had many descendants, suggesting these genotypes had been circulating for enough time to generate mutations. Isolates from the Beijing family (node 6, SIT 1) seemed to have evolved independently for the largest component in the spoligoforest.

### Descriptive statistics of clustering of mycobacterial isolates

Out of the 400 isolates, 365 strains (91%) were grouped in 36 clusters (range: 2-92). There were 12 clusters (33%) with size 2, 19 clusters with size 3-19 (53%) and 5 clusters with size ≥20 (14%) (Figure 3). Averaging across people the median cluster size was 25 (IQR 9-92). The proportion of clustering was similar in the HIV-population (93%) as compared with the patients with a HIV negative/unknown status (90%).

**Figure 3.**
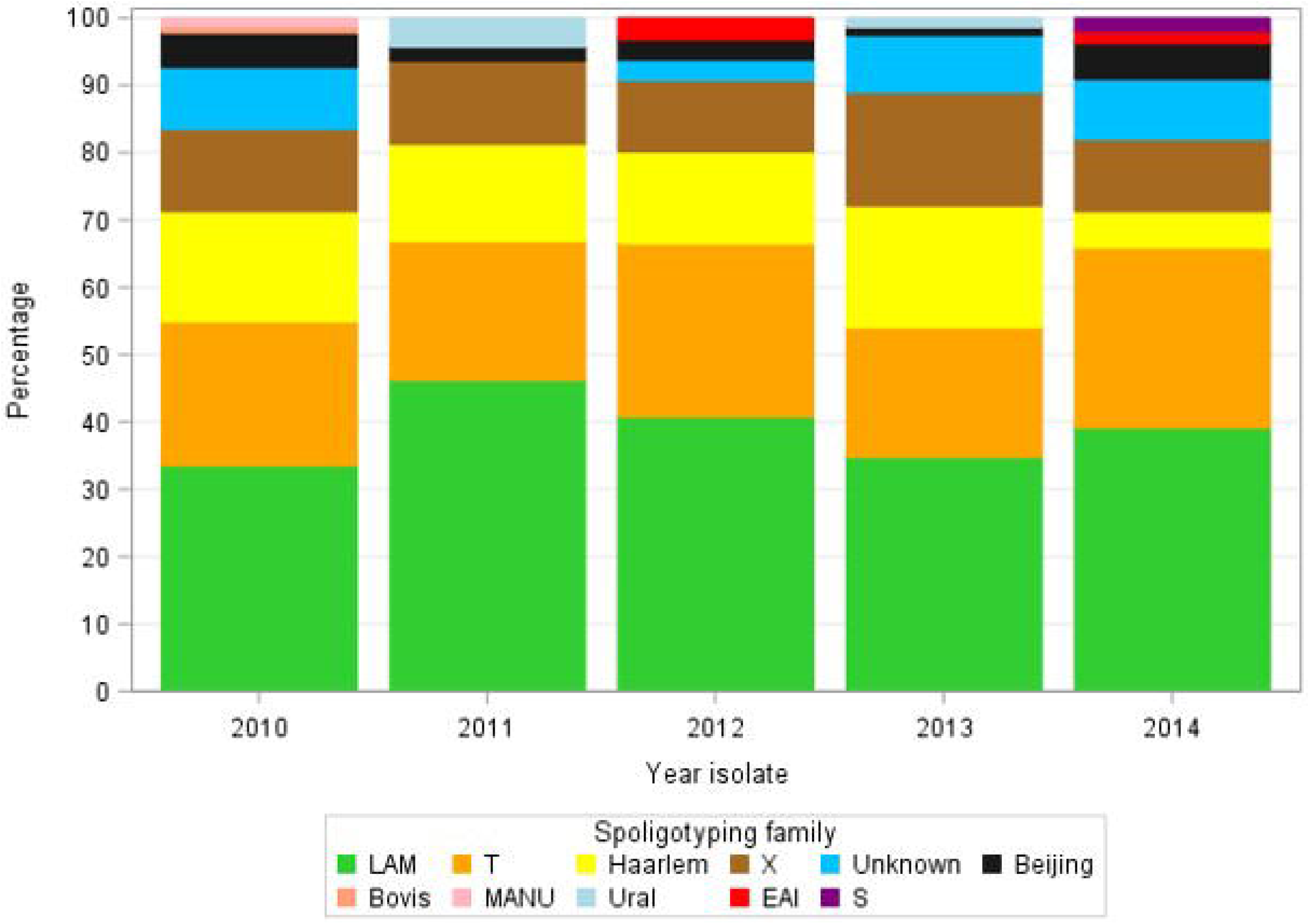
Cluster size distribution among the patients with clustered mycobacterial strains isolated from 2010-2014. In the X axis, the cluster size is given. The Y axis represent the frequency of the given cluster size. The range of the cluster size was 2-92 isolates.

### Estimation of the proportion of TB cases due to recent transmission

The proportion of tuberculosis cases due to recent transmission (RTI_n-1_) by the traditional ‘n-1’ method was 82% in the five years of the study. This proportion minimally decreased when using different time windows, 78% for the 2-year window, 80% for the 3-year window and 81% for the 4-year window (Table 3).

**Table 3.**
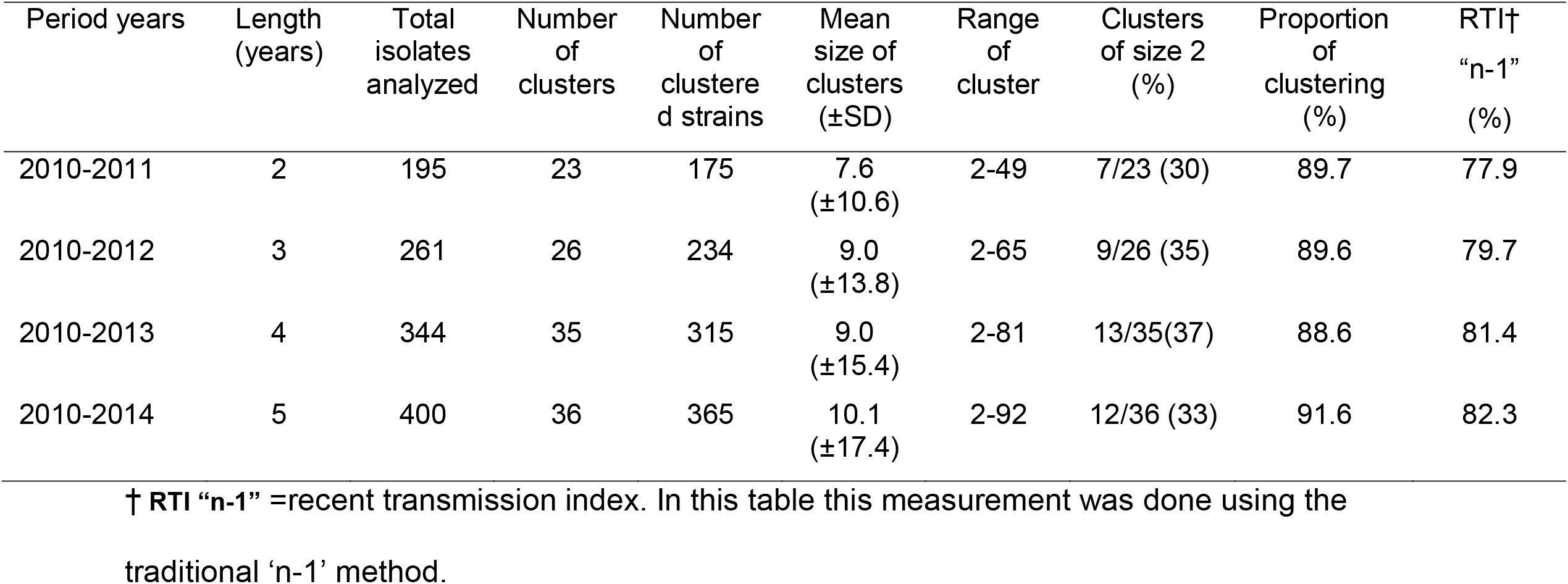
Descriptive statistics comparing clustering of mycobacterial isolates depending years of isolation.

In the sensitivity analysis conducted after exclusion of clusters with ≥ 20 isolates, the proportion of clustering was 81% (146/181) and the RTI_n-1_ was 64%.

### Independent factors associated with clustering in the HIV-infected individual

Out of the 132 patients with HIV confirmation, we were able to collect the majority of demographic, behavioral and clinical variables for 113 patients. These cases were primarily men (76%), with a median age of 36 years (IQR 28-45). At the time of the tuberculosis diagnosis, 65% where at HIV clinical stage 3, and 63% had a CD4 count of less than 200 cells/mm^3^. The proportion of clustering in the isolates of this sub-group was 93% (105/113) similar to the proportion in the whole HIV-population.

In bivariate analysis, of all the demographic variables investigated (Table S4), only gender of the patient seemed to be of relevance. Female patients had a proportion of clustering of 100%, higher than the one detected in male patients (91%, OR=6.0, 95% CI 0.3-112.2).

Patients that submitted pulmonary samples had clustered isolates in 97% of the cases, whereas patients who submitted extra-pulmonary samples had a lower proportion (85%, OR=4.3, 95% CI 1.0-17.7). Similarly, patients with a positive smear result had a higher proportion of clustering (100%) than patients with a negative smear result (89%, OR=11.1, 95% CI 0.6-206.3). We also noted that patients with no self-reported alcohol consumption had higher proportion of clustered isolates (100%) than patients with alcohol consumption (92%) but these findings did not reach statistical significance (OR=7.0, 95% CI, 95% CI 0.4,136.6). Previous history of tuberculosis, prison, drug use and tobacco consumption were not associated with the proportion of clustering (Table S5).

## DISCUSSION

We uncovered a high level of recent transmission among tuberculosis cases from Guatemala, Central America between 2010-2014. The proportion of clustered cases detected in this study, 91% was slightly higher to other spoligotyping-based studies reported among tuberculosis cases in the South of Mexico (78%) and in Honduras (84%), both neighboring areas of Guatemala [19, 20].

A systematic review analyzing data from 36 studies conducted in 17 countries estimated the median tuberculosis clustering proportion at 39%, but a wide range was detected (7-72%) [4]. In those studies, IS6110-RFLP was the main method for genotyping, in contrast of our use of spoligotyping. It has been shown that spoligotyping, owed to its low-resolution power, can overestimate the number of clustered isolates by up to 50% [21]. If that was the case, the proportion of clustering in this study might be at least of 45%, still suggesting a high rate of transmission.

We did not find differences in the level of recent transmission using different time windows. Moreover, the proportion of clustered isolates did not vary according to HIV status. HIV status has been reported to be of importance exclusively in patients ≥45 years of age [16]. We did not observe this association in our results, probably owed to the high levels of recent transmission in the whole population.

We found that risk factors for clustering in the HIV population were pulmonary tuberculosis and having smear-positive tuberculosis. Our findings resemble the ones obtained in other molecular studies [22-24]. In San Francisco, US cervical lymphatic tuberculosis and non-respiratory tuberculosis were associated with low clustering proportion (adjusted odds ratio 0.55, 95% CI 0.31–0.96, and 0.55, 95% CI 0.37–0.83 respectively) [23]. Similar results were reported in Houston, US, where 65% of patients with pulmonary tuberculosis had clustered strains in contrast with 58% of patients with extra-pulmonary tuberculosis [22]. Moreover, in a systematic review of risk factors associated with recent transmission of tuberculosis, sputum smear positivity was identified as one of them (pooled odds ratio 1.4, 95% CI 1.2-1.6) [25]. These results suggest that HIV-infected cases with a pulmonary tuberculosis and/or smear-positive tuberculosis are part of recent transmission chains, confirming previous findings showing that HIV-infected tuberculosis cases are equal infectious than HIV-seronegative cases when they are smear-positive [26].

We found evidence of mixed infection in two patients who were infected with different strains in their concurrent pulmonary and extra-pulmonary samples. The proportion of mixed infections in tuberculosis is still unclear, the limited evidence shows proportions from 8% to 57% in high burden areas [27]. A previous report from Uganda shown that 26/51 (51%) of HIV-infected individuals with *M. tuberculosis* strains in both sputum and blood had discordant genotypes [28]. Low CD4 T-cell count has been associated with the presence of mixed tuberculosis infections [29], a feature present in the HIV-infected individual with a mixed infection from our study. These mixed infections might be the result of a single transmission event, multiple transmission events or if a second infection causes the relapse of the first infection [30]. There is an urgent need to further our research in this areas as mixed infections can impact strategies aiming to control and treat tuberculosis [31]

As a second aim we described for the first time, the molecular epidemiology of tuberculosis in Guatemala. The most common spoligotypes families found in this study, LAM, T and Haarlem are similar to the ones obtained in South America, although their proportion varied according to the country as recently reviewed [32, 33]. Nevertheless, the X family accounted for less than 7% in the majority of these studies and we found a prevalence of 13%. In Honduras, the only Central American country with genotyping studies, LAM family accounted for 55% of the isolates, followed by Haarlem (16%), T (16%) and X (6%) [20].

The most prevalent genotypes identified in this study belong to the Euro-American phylogenetic lineage 4, according to a genotype classification recently published [34]. Lineage 4 is one of the seven lineages in which are categorized *M. tuberculosis* complex strains and is the one with the largest distribution in the world [35, 36]. Its introduction in the Americas is considered to be owed to the European colonization of the continent after the 1500s [33].

Prior to this work, only 21 isolates from Guatemala have been documented in the international spoligotype SITVIT2 database, which currently comprises 103, 856 strains with 9, 658 patterns (http://www.pasteur-guadeloupe.fr:8081/SITVIT2/). The addition of the spoligotype-patterns from the study samples will increase our knowledge of the circulating genotypes in Central America (submitted to SITVIT2 on October 2019). Moreover, the detection of a cluster of nine isolates that previously has been reported just as an orphan isolate in the US in 2006, from the Harlem family, requires further research to understand its transmission dynamics.

This study has several important limitations. We were not able to use an additional second genotyping to better discriminate our spoligotypes, which more likely resulted in an overestimation of the level of transmission. To confirm our results a second genotyping method such as mycobacterial interspersed repetitive-unit–variable-number tandem-repeat (MIRU-VNTR) typing or whole genome sequencing (WGS) is warranted [37]. Second, our study population was restricted to one health center so ascertainment bias could have occurred [38], limiting the generalizability of our results. Third, we utilized a convenience sampling with a low sampling rate, which understates the level of transmission [12, 39, 40]. Fourth, information bias is possible owed to the quality of our data [38]. Several of the potential risk factors for clustering were collected months of years before the tuberculosis diagnosis or they had incomplete information on the level of exposure. Nevertheless, if this is bias is present is more likely a non-differential misclassification bias. Based on these limitations, we believe our results are exploratory in nature but still suggest high levels of transmission in Guatemala and might explain the lack of reduction of tuberculosis in the last ten years in the country. For example, a recent molecular study, using whole genome sequencing identified an outbreak of the Beijing family in a poor area in a neighborhood in Guatemala City close to a prison center [41] for at least 2 years, showcasing the risk of outbreaks that might last a long time and the ongoing transmission in the community.

In conclusion, there might be high levels of ongoing transmission of *M. tuberculosis* in Guatemala as indicated by clustering in a convenience sample. Among HIV-infected patients, clustering was more likely in pulmonary disease. Further prospective studies in Guatemala and neighboring countries with novel genotyping techniques and larger sampling fractions are urgently needed to further characterize the molecular diversity and transmission dynamics of tuberculosis in the Central American region.

## Data Availability

Data will be available under request.

## Acknowledgements

MEC was supported by the Schlumberger Foundation Faculty, Houston, TX, USA, for the Future Fellowship. Thanks to all the staff and personnel from ASI, HIV clinic ‘Clínica Familiar Luis Ángel García’/Hospital General San Juan de Dios, particularly Brenda Guzman and Haydee Ortíz. Many thanks to Dr. Nalin Rastogi and Dr. David Couvin from the Institut Pasteur in Guadeloupe for the support and guidance with the use of SpolSimiliartySearch and to Dr. Zach Aandahl and Dr. Mark Tanaka from the University of New South Wales for the technical support using the MERCAT package.

